# Effects of environmental polycyclic aromatic hydrocarbon exposure and pro-Inflammatory activity on Type 2 Diabetes Mellitus

**DOI:** 10.1101/2021.10.08.21264766

**Authors:** Shweta Srivastava

## Abstract

**Background:** Polycyclic aromatic hydrocarbons (PAHs) are formed due to incomplete combustion and known for their potential impact and persistence in the environment. PAHs exposure have been linked to cause adverse health effect including cancer and genetic mutations. The understanding of metabolic effects of PAH exposure are still less clear especially in the presence of pro-inflammatory stress like alcoholism or diabetes.

**Objective:** The aim of this article is to understand the metabolic effects of PAH exposure by analyzing the clinical biomarkers. This study has also accessed the interactive impact of PAH and other proinflammatory factors, like alcohol intake on the metabolic syndrome, especially Type 2 Diabetes Mellitus (T2DM).

**Methods:** All the data in this study are retrieved from CDC NHANES (2015-16). We investigated urinary levels of hydroxylated PAH metabolites (OH-PAHs) along with demographic, clinical and laboratory data. Questionnaire data for alcohol use and diabetes status were also included along with laboratory data. Laboratory measures included in the study were levels of PAHs, glycohemoglobin, glucose, cholesterol, lipids, triglyceride, complete blood count, lymphocytes, and monocytes. Generalize linear model Univariate factorial ANOVA was used to evaluate the group differences (both between the groups; as well as across all the groups) in the demographics, PAH exposure, drinking patterns, clinical data, and biomarker levels. Linear regression model was used to analyze the association of biomarkers, PAH exposure and drinking data. Multivariable regression model was used for multi-independent model to assess comorbidity association and their effect sizes on the clinical outcomes.

**Results:** BMI (p=0.002), and age (≤0.001) are independent demographic risk factors for T2DM in high PAH exposure. Acute proinflammatory activity characterized by CRP, is augmented by elevated monocyte levels (p≤0.001) and stepwise addition of 1-HN (p=0.005), and 2-HN (p=0.001) independently. Prevalence of highest average drinks over time is observed in the high PAH exposure; with males drinking almost twice compared to females in Gr.3. Pathway response of T2DM shows sexual dimorphism; with males showing association with triglycerides (p≤0.001), and females with CRP (p=0.015) independently with HbA1C. The arrangement of CRP, absolute monocyte levels, serum triglycerides and average drinks over time predict the HbA1C levels (adjusted R^2^=0.226, p≤0.001) in individuals with high PAH exposure.

**Discussion:** In this large dataset investigation on humans, the adverse effects of high exposure of PAHs identified candidate demographic risk factors. Preclinical experimental studies on mice have suggested that PAHs exposure induces lipid metabolic disorders in a time-dependent manner, which we found in humans too. Sexual dimorphism is observed in alcohol drinking with males drinking more in the high PAH exposure group. Alcohol drinking as an independent factor associated with the DMT2 indicator, HbA1C in individuals with high PAH exposure.

**Highlights:**

1. BMI and Age are demographic risk factors for Diabetes Mellitus Type 2 (DMT2) in high PAH exposure
2. Acute proinflammatory activity characterized by CRP, is augmented by elevated monocyte levels and 1-HN and 2-HN independently
3. Prevalence of higher average drinks over time is observed with high PAH exposure
4. Pathway of DMT2 shows sexual dimorphism, with males showing association with triglycerides, and females with CRP independently with HbA1C
5. The arrangement of CRP, absolute monocyte levels, serum triglycerides and average drinks over time predict the HbA1C levels in individuals with high PAH exposure.

## INTRODUCTION

Polycyclic aromatic hydrocarbons (PAHs) are ubiquitous environmental pollutants linked to many adverse health effects [1, 2]. Polyaromatic Hydrocarbons (PAHs) are a group of chemicals that occur naturally in coal, crude oil, and gasoline. They also are produced when coal, oil, gas, wood, garbage, and tobacco are burned [3, 4]. PAH can also be present in food like charcoal grilled meat and processed food [5, 6]. National Institute for Occupational Safety and Health (NIOSH) recommends workplace air exposure levels of PAHs at less than 0.1 mg/m^3^ [7].

Upon entering the body, PAHs are metabolized and eliminated via urine. Urinary concentrations of PAH metabolites, specifically mono-hydroxylated PAHs (OH-PAHs), have been used as biomarkers of human exposure to select PAHs, including naphthalene, fluorene, phenanthrene, and pyrene [8, 9].

PAHs can form small airborne particles and cause a pronounced pulmonary inflammatory response and lung cancer [10, 11]. PAHs have been found to reach to heart through circulation causing cardiotoxicity such as atherosclerosis, cardiac hypertrophy, arrhythmias, and contractile dysfunction [12, 13]. The transport of PAHs to systemic circulation can also significantly affect hepatic metabolism [14-16] since liver is major metabolic organ of PAHs. Inhaled Benzo[a]pyrene are metabolized to generate reactive oxygen species (ROS) in the liver, directly contributing to the hepatoxicity [17].

PAHs have been found to increase likelihood of Type 2 Diabetes Mellitus (T2DM) by 78-124% [18]. This risk is confounded by the fact that some PAHs increase independent risk factors of T2DM such as obesity, alcohol intake, elevated blood pressure and blood lipid [19, 20]. However, there are gaps in the understanding of the clinical presentation or altered mechanisms that can clearly ascertain the role of PAHs on T2DM. Its adverse effects on T2DM and its clinical marker, HbA1C are still under investigation. Importantly, interaction of heavy or binge alcohol intake and PAHs on the exacerbation of metabolic syndrome is not clear.

Majority of the research done so far are mainly focused on respiratory toxicity of inhaled PAHs. In the present study, the 2015–2016 NHANES data for seven urinary metabolites of PAH exposure namely (with their code in parenthesis for NHANES tracking): 1-hydroxynaphthalene (URXP01), 2-hydroxynaphthalene (URXP02), 2-hydroxyfluorene (URXP04), 3-hydroxyfluorene (URXP03), 1-hydroxyphenanthrene (URXP06), 2- & 3-hydroxyphenanthrene (URXP25), and 1-hydroxypyrene (URXP10) was used to assess their effects on T2DM. Primary aim of this study was to characterize the exposure effect of PAHs on T2DM; and examine the modifying effects of alcohol intake, blood pro-inflammatory mediators and lipids. A model was also developed to describe the interaction of high PAHs, modifying risk factors on T2DM. Lastly, the role of demographic confounders was also explored.

## METHODOLOGY

### Study Participants, questionnaires, and examination data

Population data were taken from the 2015–2016 NHANES provided by the National Center for Health Statistics (NCHS) (https://wwwn.cdc.gov/Nchs/Nhanes/2015-2016/DEMO_I.htm). All the laboratory measurements were performed by CDC’s Division of Laboratory Sciences at the National Center for Environmental Health. Survey data were collected at the mobile examination center interview room on the day of the health exam by CDC. The demographics data extracted for the study includes information on the Survey participant’s household interview and examination status, gender, age, and pregnancy status. Examination data includes body measures and blood pressure. Questioner data has information about drinking habit and known diabetes status.

### Laboratory Data

All the laboratory methods for blood and urine samples were performed by NHANES that has been described thoroughly at: (https://wwwn.cdc.gov/nchs/nhanes/continuousnhanes/labmethods.aspx?BeginYear=2015). Laboratory measured data in the study are levels of PAHs, Glycohemoglobin, Glucose, Cholesterol, Lipids, Triglyceride, complete blood count, Lymphocytes, Monocytes, and hepatitis conditions.

### Analytical method for urinary PAH measurement

The analytical procedure followed by NHANES involves enzymatic hydrolysis of glucuronidated/ sulfated OH-PAH metabolites in urine, extraction by on-line solid phase extraction, and separation and quantification using isotope dilution high performance liquid chromatography-tandem mass spectrometry (on-line SPE-HPLC-MS/MS) [8].

### Selection of eligible participants

After data collection from the NHANES website, samples where data for any of the following were missing were excluded: missing examination status, age below 18 years, pregnancy, missing BMI, missing laboratory data for Glycohemoglobin or Poly aromatic hydrocarbons (PAHs). Individuals with missing alcohol drinking data for past 12 months were also excluded.

The specific analytes measured in this method are OH-PAHs, namely URXP01, URXP02, URXP03, URXP04, URXP06, URXP25, and URXP10 (In the NHANES database). PAH metabolites data below lower limit of detection (LLOD) of the analytical method levels were excluded (Fig. 1).

**Figure 1.**
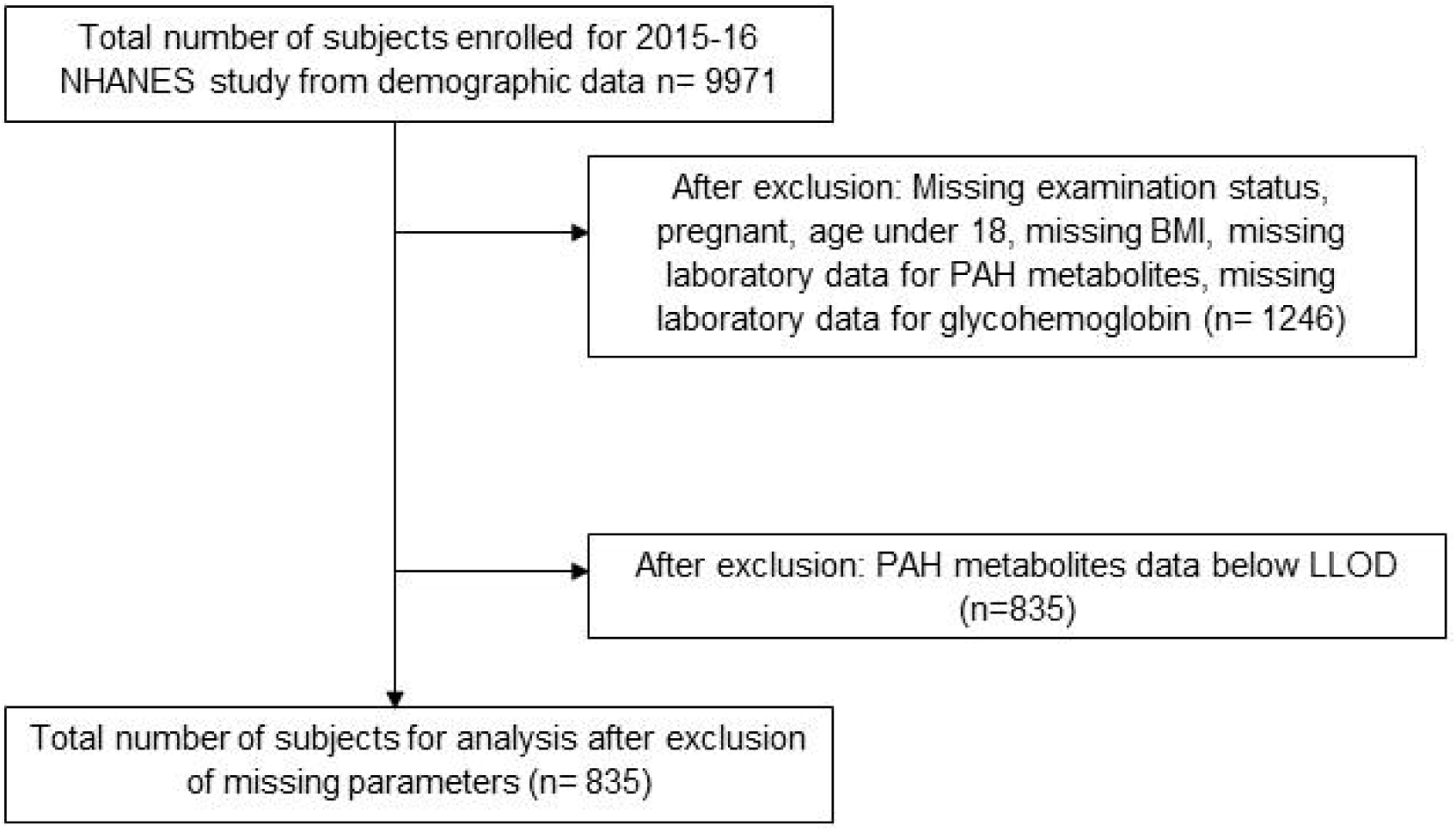
Flow Chart illustrating the selection of eligible participants. Eligible participants and those included in the analysis for the associations between urinary PAH levels and other clinical parameters.

### Statistics

Participants were categorized for this investigation based on their PAH metabolite exposure range as per NHANES data (Supplement Table 1). Population based on each metabolite levels was categorized into 3 groups: 1 below geometric mean exposure range, 2 geometric mean exposure range and 3 higher than geometric mean exposure of the reference range of URXP01 calculated by NHANEs (<u>Fourth National Report on Human Exposure to Environmental Chemicals Update</u> (cdc.gov)). In this study, we used the exposure levels of 1-HN (stated as URXP01 in the NHANES database) for the analyses purpose.

Male with 14≥ drinks/ week, and females with 8≥ drinks/ week are heavy drinkers as per NIAAA classification (<u>Drinking Levels Defined | National Institute on Alcohol Abuse and Alcoholism (NIAAA</u>) (nih.gov). Low risk or moderate drinking are who drank 2≤ drinks/ day as males, and females as 1≤ drinks/ day (<u>Drinking too much alcohol can harm your health. Learn the facts | CDC</u>).

Univariate factorial ANOVA was used to evaluate the group differences (both between the groups; as well as across all the groups) in the demographics, PAH exposure, drinking patterns, clinical data, and biomarker levels in context. Linear regression model was used to analyze the association of biomarkers and clinical, PAH exposure and drinking data. Demographic data were used as covariates, if found to be significantly different in the between-group analyses. Multivariable regression model was used for multi-independent model to assess comorbidity association and their effect sizes on the clinical outcomes. IBM SPSS version 27 (IBM®, Armonk, New York USA) was used to compute all the statistical analysis and correlations in the study. Microsoft 365 version (Microsoft Corp., Redmond, Washington USA) was used for data processing and Table/s development. Figures were processed using GraphPad Prism (GraphPad, San Diego, CA USA). Data in table/s are presented as Mean and standard deviation (M±SD). Statistical significance was set at p < 0.05.

## RESULTS

### Demographics, Drinking Profile and Diabetic Indexes by PAH Exposure Level

Age was significantly higher in Gr. 3 compared to Gr. 1 (Table 1). Notably, all the individuals in each group showed overweight to borderline obese, especially Gr. 1 and Gr. 2. Gr. 3 individuals drank on an average daily more than the rest of the two group individuals, and significantly higher (p=0.015) compared to the Gr. 1 (Table 1).

**TABLE 1.** Demographic, drinking, PAH, Lipids, blood cell values, C-reactive protein and diabetic markers are tabulated. Statistical significance was set at p≤0.05. Data presented as Mean±SD.

| Measures | Group 1 (low) 1-Hydroxynaphthalene |  |  | Group 2 (Normal) 1-Hydroxynaphthalene |  |  | Group 3 (High) 1-Hydroxynaphthalene |  |  | *all groups p-value* |
| --- | --- | --- | --- | --- | --- | --- | --- | --- | --- | --- |
|  | Males (n= 287; %) | Females (n= 218; %) | Total (n = 505) | Males (n= 6; %) | Females (n= 7; %) | Total (n= 13) | Males (n= 186; %) | Females (n= 131; %) | Total (n= 317) |  |
| Age (years) <sup>c</sup> | 45.57 ± 17.76 | 43.46 ± 17.58 | 44.66 ± 17.69 | 43.17 ± 22.41 | 44.71 ± 13.22 | 44.00 ± 17.24 | 49.94 ± 16.52 | 48.98 ± 15.88 | 49.54 ± 16.24 | NS |
| BMI (kg/m<sup>2</sup>) <sup>b#, c</sup> | 29.88 ± 6.33 | 31.507 ± 8.69 | 30.584 ± 7.48 | 29.23 ± 8.05 | 34.00 ± 7.37 | 31.80 ± 7.76 | 27.55 ± 5.66 | 29.83 ± 7.31 | 28.50 ± 6.48 | ≤0.001 |
| Average Drink per Day in past 1 year | 3.21±2.71 | 2.09±1.42 | 2.74±2.32 | 3±1.67 | 1.86±.69 | 2.38±1.32 | 3.87± 2.80 | 2.21±1.68 | 3.22±2.65 | ≤0.037 |
| PAH metabolite level (ng/gm of creatinine) |  |  |  |  |  |  |  |  |  |  |
| 1-hydroxynaphthalene (ng/g of creatinine) <sup>**</sup> | 1164.49 ± 788.29 | 1098.06 ±750.47 | 1135.81±772.14 | 3726.81±147.24 | 3760.83±180.34 | 3745.13±160.02 | 52465.91±329582.65 | 165450.57±845589.68 | 99156.73±600710.65 | NA |
| 2-hydroxynaphthalene (ng/g of creatinine) <sup>c</sup> | 5893.71±7543.00 | 7866.85±6303.15 | 6745.48±7095.74 | 5561.87±2065.07 | 8140.95±2963.36 | 6950.61±2821.07 | 13022.58±8308.54 | 18245.61±13760.66 | 15180.99±11178.08 | ≤0.001 |
| 2-hydroxyfluorene (ng/g of creatinine) <sup>a, b#, c</sup> | 234.74±349.28 | 225.86±279.47 | 230.91±320.74 | 589.49±226.21 | 336.84±329.75 | 453.45±304.75 | 1169.67±1609.27 | 1184.38±1138.77 | 1175.75±1431.68 | ≤0.001 |
| 3-hydroxyfluorene (ng/g of creatinine) <sup>a, b, c</sup> | 105.74±116.42 | 95.19±145.33 | 101.18±129.66 | 279.80±89.09 | 212.44±260.72 | 243.53±196.25 | 730.24±761.15 | 786.40±830.78 | 753.45±789.87 | ≤0.001 |
| 1-hydroxyphenanthrene (ng/g of creatinine) <sup>a#, c</sup> | 129.40±123.75 | 135.78±117.36 | 132.15±120.96 | 286.67±277.73 | 117.16±58.92 | 195.40±203.99 | 278.06±528.98 | 278.04±294.85 | 278.05±446.74 | ≤0.001 |
| 2- & 3-hydroxyphenanthrene (ng/g of creatinine) <sup>c</sup> | 168.42±273.48 | 144.62±93.45 | 158.15±215.27 | 370.29±401.85 | 153.63±69.61 | 253.63±286.96 | 473.01±880.40 | 422.63±614.16 | 452.19±780.75 | ≤0.001 |
| 1-hydroxypyrene (ng/g of creatinine) <sup>a, c</sup> | 137.14±148.64 | 153.18±100.39 | 144.07±130.16 | 537.42±693.53 | 131.80±77.49 | 319.01±497.71 | 419.58±602.28 | 415.68±461.92 | 417.97±547.85 | ≤0.001 |
| Cholesterol and Triglycerides |  |  |  |  |  |  |  |  |  |  |
| Total Cholesterol (mg/dL) | 188.11±43.81 | 183.94±38.79 | 186.33±41.75 | 171.83±54.52 | 193.86±48.22 | 183.69±50.31 | 187.70±40.62 | 192.67±39.51 | 189.76±40.18 | NS |
| HDL (mg/dL) <sup>c</sup> | 47.79±14.64 | 55.23±16.92 | 50.96±16.06 | 45.17±11.50 | 61.71±14.03 | 54.08±15.07 | 51.45±19.34 | 57.33±16.62 | 53.88±18.47 | 0.061<sup>#</sup> |
| LDL (mg/dL) <sup>c</sup> | 112.30±35.54 | 105.86±30.42 | 109.59±33.56 | 88.50±72.83 | 132.67±27.57 | 115.00±47.87 | 115.67±39.88 | 112.16±32.13 | 114.36±37.08 | NS |
| Triglycerides (mg/dL) <sup>a#</sup> | 106.16 ±63.16 | 99.82 ±67.97 | 103.48 ±65.15 | 201.50 ±118.09 | 132.33 ±63.71 | 160.00 ±83.37 | 129.01 ±103.99 | 108.10 ±54.03 | 121.25 ±89.12 | 0.045 |
| Candidate/ Target Blood Cell Measures |  |  |  |  |  |  |  |  |  |  |
| Lymphocyte Absolute number (1000 cells/uL) <sup>c</sup> | 2.07±0.66 | 2.40±0.85 | 2.21 ±0.76 | 2.02±0.47 | 2.09±0.39 | 2.06±0.41 | 2.23±0.77 | 2.53±0.75 | 2.35±0.77 | 0.015 |
| Monocyte Absolute number (1000 cells/uL) <sup>c</sup> | 0.60±0.20 | 0.57±0.18 | 0.59±0.19 | 0.66±0.38 | 0.54±0.10 | 0.59±0.25 | 0.63±0.19 | 0.61±0.20 | 0.62±0.19 | 0.071 |
| Blood Glucose and diabetes measures |  |  |  |  |  |  |  |  |  |  |
| Glycohemoglobin (%) <sup>c#</sup> | 5.74±1.07 | 5.71±1.12 | 5.73±1.09 | 5.45±0.73 | 6.23±1.75 | 5.87±1.38 | 5.92±1.16 | 5.78±0.97 | 5.86±1.09 | NS |
| Glucose, refrigerated serum (mg/dL) | 104.96±43.22 | 99.57±32.45 | 102.66±39.06 | 108.17±31.19 | 94.33±12.08 | 101.25±23.68 | 107.67±47.28 | 101.98±38.97 | 105.32±44.06 | NS |
| Inflammation markers |  |  |  |  |  |  |  |  |  |  |
| HS C-Reactive Protein (mg/L) <sup>c#</sup> | 2.93±5.17 | 5.35±7.08 | 3.96±6.17 | 5.58±5.21 | 3.31±5.41 | 4.45±5.20 | 4.82±10.72 | 5.30±7.54 | 5.02±9.53 | NS |

Footnote: BMI: Body Mass Index. HDL: High density Lipoprotein. LDL: Low Density Lipoprotein. <sup>a</sup> Significant difference between group 1 and 2. <sup>b</sup> Significant difference between group 2 and 3. <sup>c</sup> Significant difference between group 1 and 3. # Trend level of significance. <sup>a</sup>~, <sup>b</sup>~, <sup>c</sup>~ #: Trend level of significant difference between two groups. NS: Not Significant. NA: Not Applicable. \*\* comparisons were not shown since the groups were distributed by the levels of 1-hydroxynapthalene, thus the differences were all significant.

### Association of Diabetic Indexes and demographic and drinking Profile by PAH Exposure

HbA1C and age were very significantly associated (R=0.331, p≤0.001) (Fig. 2a) in Gr. 3 (high exposure group of 1-Hydroxynapthelene). HbA1C and BMI were also very significantly (p=0.002) associated (R=0.230) (Fig. 2b) in Gr.3. In this univariate regression model, diabetic index (HbA1C) as a dependent variable showed unique relationship with candidate demographic indices exclusively in the high exposure group of 1-Hydroxynapthelene.

**Figure 2.**
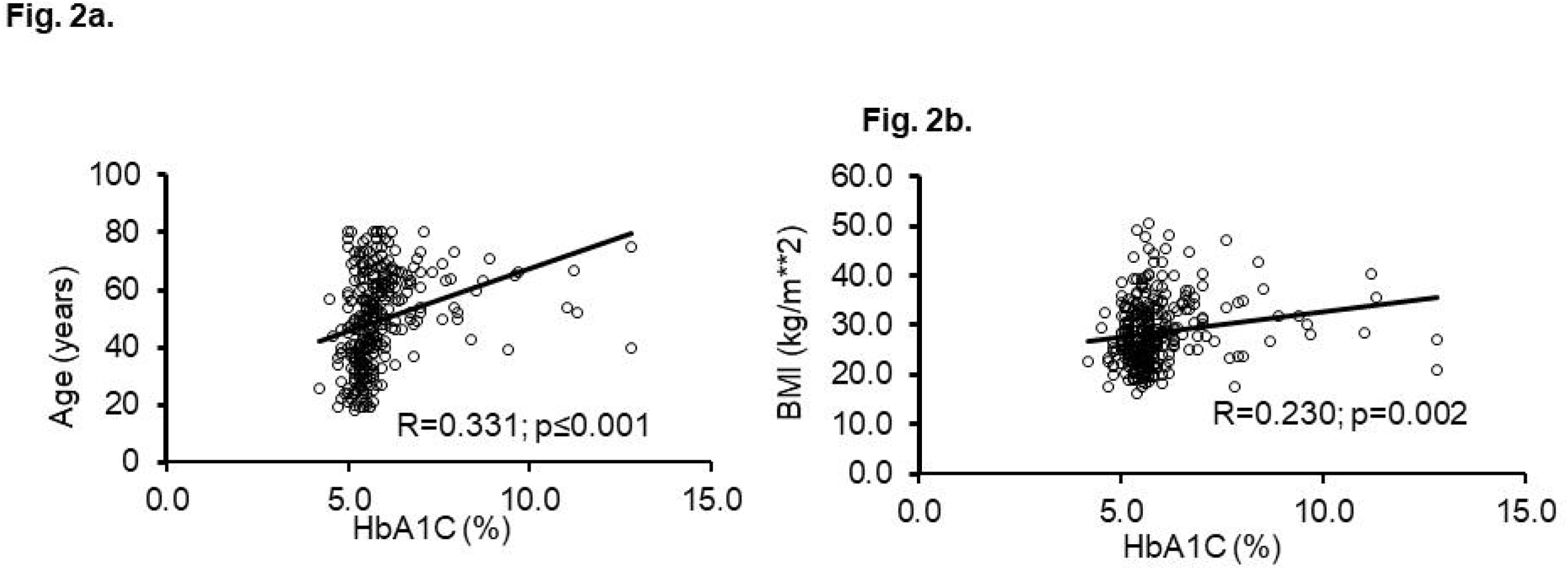
Association of HbA1C and demographic parameters in high exposure group 3 of the study. Fig. 2a: Association of HbA1C and age. Fig. 2b: Association of HbA1C and Body Mass Index (BMI). Raw data plotted on x- and y-axes. Effect sizes/correction index is noted along with the p-value. Statistical significance was set at p≤0.05.

### Association of Inflammatory Markers and PAH

Univariate analysis of regression showed significant prediction (R=0.141, p=0.001) of C-Reactive protein by the absolute Monocyte level in Gr. 3. Multivariable independent variable regression model was used to include the contributing role of candidate PAHs that potentially are involved in the inflammation. CRP, marker of acute pro-inflammation, as a dependent variable and absolute monocyte count (or numbers) (AMC) as independent variable showed augmented effects of association with the candidate PAHs; 1-HN (R=0.142, p=0.005) (Fig. 3a), 2-HN (R=0.149, p=0.001) (Fig. 3b); respectively.

**Figure 3.**
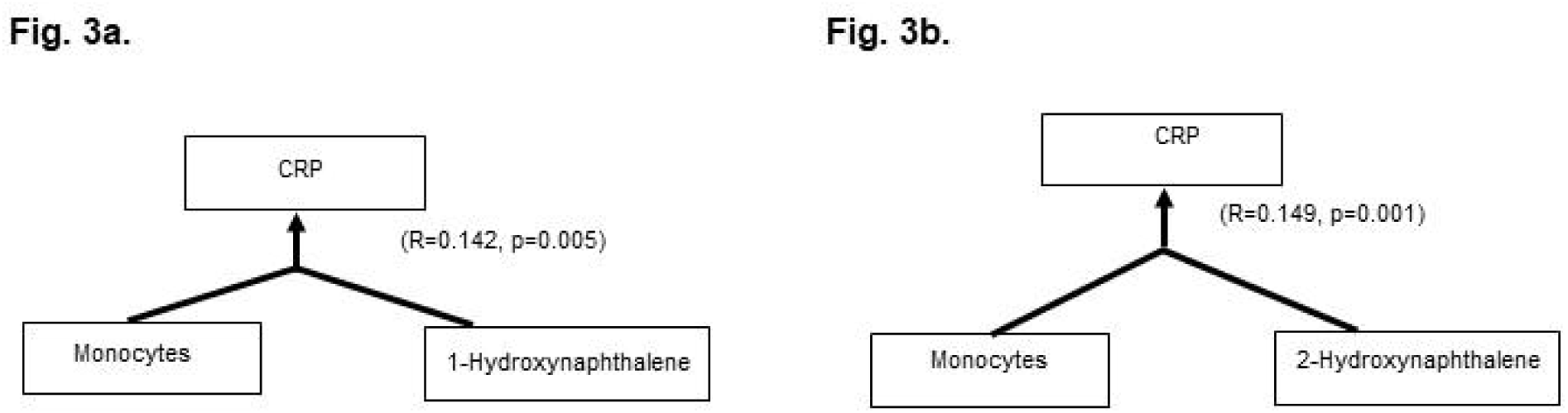
Association of C-Reactive Protein with absolute monocyte numbers in Group 3 of highly exposed subjects in multivariable regression model with PAH. 3a. 1-Hydroxynaphthalene. 3b. 2-Hydroxynaphthalene. Effect sizes/correction index is noted along with the p-value. Statistical significance was set at p≤0.05.

To estimate the role of alcohol drinking, which is also a cause of inflammation, we reviewed the levels of “Average Drinks Per Day” (ADPD) in all the groups (Fig. 4a). We found that Gr. 3 individuals tend to drink more than the rest two groups. In Gr. 3, males drank significantly higher than the females (Fig. 4b). Gr. 3 males showed significant association of HbA1C and triglycerides (Fig. 4c). On the other hand, Gr. 3 females showed significant association of HbA1C and CRP (Fig. 4d).

**Figure 4.**
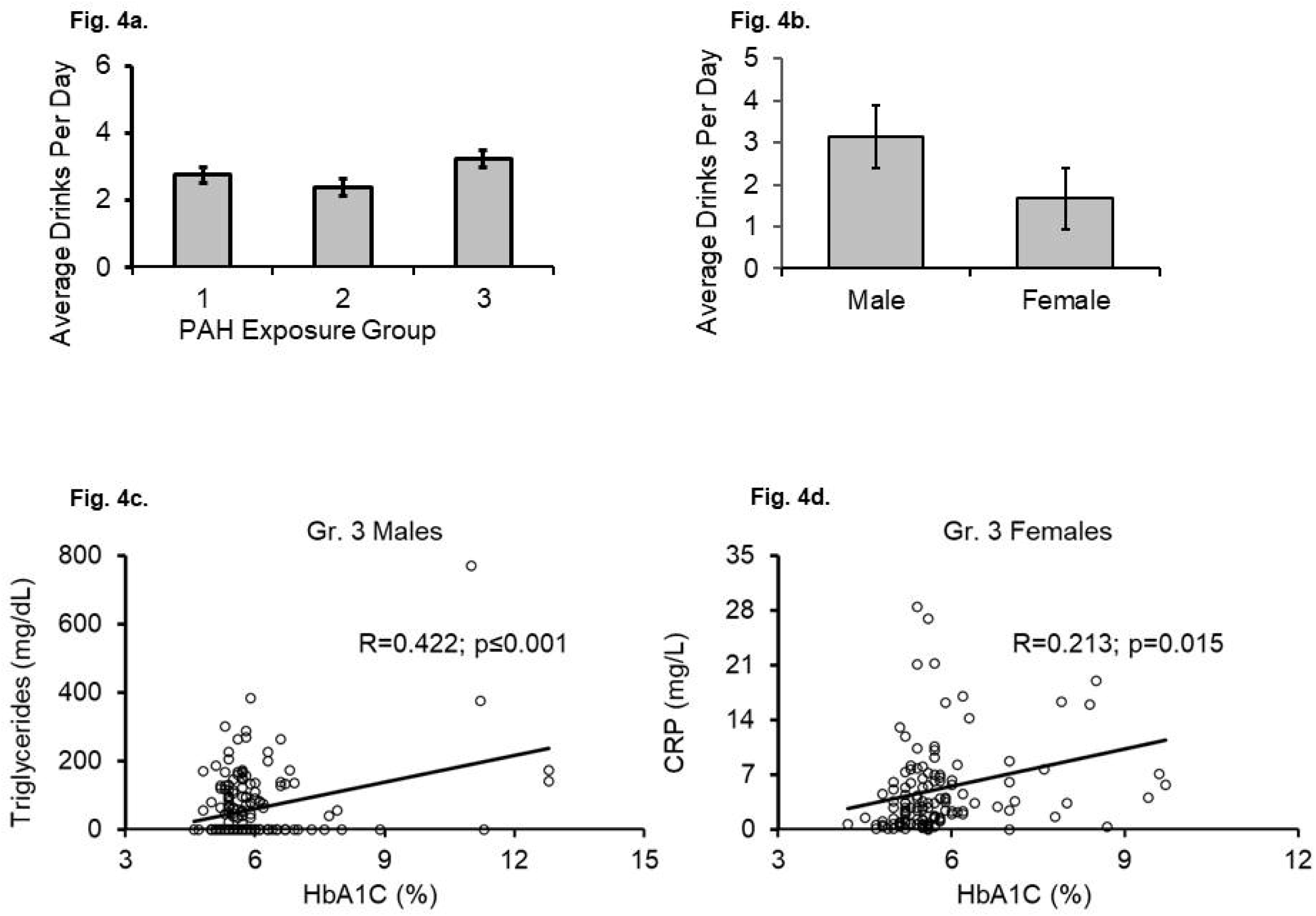
Alcohol drinking and PAH exposure; and interaction of diabetes and pro-inflammatory markers as sexual dimorphism in Gr. 3. Fig. 4a: Gr. 3 individuals drank more than the other groups. Fig. 4b: Males drank more than the females in Gr. 3. Gr. 3: Association of triglycerides and HbA1C in Gr. 3 males. Fig. 4d: Association of CRP and HbA1C in Gr. 3 females. Group 1: below geometric mean exposure, group 2: geometric mean exposure and group 3: higher than geometric mean exposure of the reference range. Statistical significance was set at p<0/05. Data presented as Mean ± Standard Error.

With this finding on inflammation and immune activity, we evaluated the overall impact on the Diabetic index, HbA1C in context of CRP, candidate lipid/s, drinking measure/s and immune activity in the Gr. 3. HbA1C was significantly predictable with CRP and AMC (Fig. 5), although CRP by itself showed only a close to trend level of significance for direct relationship with HbA1C. This arrangement further augmented its effects with simultaneously increasing order of statistical significance in a stepwise manner with Triglycerides and Average Drinks Per Day were included respectively as added independent variable/s in this multivariable prediction model.

**Figure 5.**
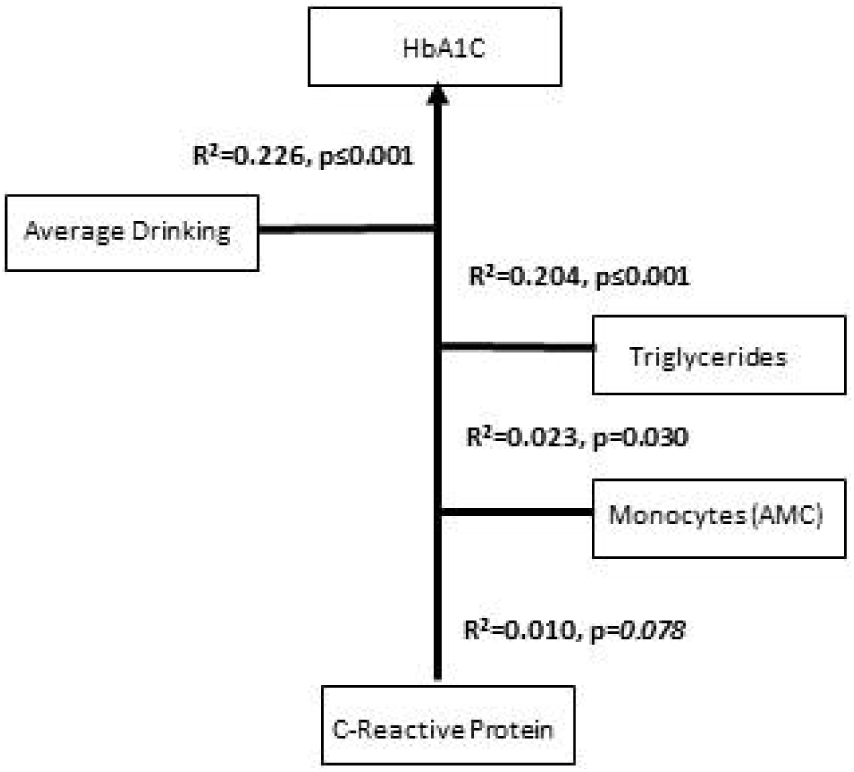
Stepwise multivariable regression analysis model illustrates the increasing effects on HbA1C as diabetic index by the pro-inflammatory activity presented as CRP, absolute monocyte levels, serum triglyceride along with average drinking per day in the Gr. 3. Statistical significance was set at p<0/05. R^2^ is noted along with the p-values.

## DISCUSSION

Findings from this investigation support the pathological role of the high exposure of PAH in the metabolic disorders; such as T2DM. Human studies have been performed earlier [21], and this study assessed the proinflammatory status and factors, risk factors and level of exposure to extend the present knowledge in a very large population. The focus of this study was 1-HN (1-hydroxynaphthalene, stated as URXP01 in NHANES database) and individuals exposed to this PAH yielded some noteworthy results.

Age and BMI stood out as the demographic measures that showed propensity with T2DM diabetic index HbA1C in individuals with high PAH exposure of 1-HN, while individuals with normal or low level of exposure did not. Similar study has been reported for PAH exposure, obesity for the risk assessment of T2DM in patients with lung disease [22].

Some studies report a close association of inflammation and PAH exposure [23] as well in the 2003-2008 wave of NHANES database [24]. We found similar assessment in the 2015-2015 database of NHANES database, especially in the context of the 1-HN PAH, which was the scope of this study. CRP and inflammation associated blood cells (absolute monocyte count) showed a significant relationship in individuals with high exposure levels in this study. Interestingly, inflammation and role of PHAs along with the oxidative stress has also been investigated in adolescents and pregnant women[24, 25].

One of the noteworthy findings in this study was that the individuals with high 1-HN exposure drank the most compared to the other exposure groups (normal/low). Males with high exposure of 1-HN had higher levels of average drinking than their female counterparts suggesting trends of sexual dimorphism. Almost a 2-fold higher average drinking in high PAH exposed males might be uniquely worrisome for a possibly unidentified role of PAH. Males having more average drinks is aligned with the findings of the NAS [26].

Earlier studies on mice have suggested that PAHs exposure induces lipid associated metabolic disorders in a time-dependent manner [27]. Similarly, CRP level observed in new-onset DM people diagnosed with HbA1c criterion has been recently reported [28]. However, there has always been a gap in how to address such findings in humans in context of PAH exposure and other risk factors. In this study, males with high 1-HN exposure showed close association of HbA1C and triglycerides, suggesting sex-specific exposure associated response to metabolic conditions [29]. Females with high 1-HN exposure on the other hand showed that HbA1C was closely associated with pro-inflammatory activity (CRP), suggesting propensity to the risks of chronic conditions [30]. Such a sexual dimorphism in high PAH exposure is a healthcare concern for metabolic disorder that needs more extensive evaluation; both mechanistically focus on biological pathways as well clinically with well-developed research questions.

In this study, several important clinical and biological measures involved in pro-inflammatory, pathological activity were observed to be higher as well as connected to each other. Thus, it was important to know their overall effect on the HbA1C levels in individuals with high PAH exposure [21, 31]. Indeed, the effect size on the HbA1C levels grew in a stepwise manner in context of candidate markers with potential role in inflammation and injury. As described in the multivariable model, role of several markers (representing several pathways that can be affected) on HbA1C, support evaluating PAH exposed individuals with comprehensive clinical assessment and varied tests to understand the complexity involved [32]. A study on a larger subject size such as this one, and its results may advance the understanding in the scientific literature on the adverse and harmful effects of PAH exposure and its interaction with the risk factors that are attributed to the metabolic disorders.

None-the-less, this study has several limitations that also need to be discussed. This is a single timepoint observational study, and assessment of longitudinal data is not in the scope of this investigation. No biological mechanism was tested in an experimental model in this evaluation to validate the corresponding clinical observations. Thus, no causations/ etiologies were identified or were the aims in this study. Risk factors, like BMI and age were identified in high exposure group of individuals. Aging and obesity associated with PAH exposure could be the important newer directions of research that can offshoot from the findings of this study. The focus of this study was limited to one PAH, 1-HN. However other PAHs are equally important to investigate and can also be evaluated as future directions as independent projects. This study also did not evaluate any other metabolic conditions.

In conclusion, this study identifies that the BMI and age likely are candidate demographic risk factors for Diabetes Mellitus Type 2 (T2DM) in individuals with high PAH exposure. Acute proinflammatory activity characterized by C-Reactive Protein, an acute pro-inflammatory marker, is augmented when elevated monocyte levels are observed in relevance of and 1-HN; and 2-HN independently. Prevalence of increased average drinks over time is also observed with high PAH exposure, especially in males. With high PAH exposure, T2DM shows sexual dimorphism. Males exposed with higher levels of PAH show association with triglycerides, whereas females with CRP independently predicted HbA1C. Importantly, the levels of CRP, absolute monocyte levels (suggesting its activation), serum triglyceride levels, and average alcoholic drinks predict the HbA1C levels robustly in individuals with high PAH exposure.

## Data Availability

All data produced are available online at
https://www.cdc.gov/nchs/nhanes/index.htm

## REPRINT REQUEST

Shweta Srivastava Ph.D., M.Sc.; Delia Baxter II, Rm. 428, Environment Health Institute; 580 S. Preston Street, University of Louisville; Louisville KY 40202.

## Abbreviations

AUD: alcohol use disorder
BMI: body mass index
CS: clinically significant
HbA1C: Hemoglobin A1c or Glycated hemoglobin
HD: heavy drinkers
HOMA: homeostatic model assessment for Types 2 Diabetes
TChol: total cholesterol
HDL: high density lipids
LDL: low density lipids
MD: moderate drinkers
PC: Polyaromatic Compounds
SD: social drinkers
SEQN: Respondent sequence number
RIDSTATR: Interview/Examination status
RIAGENDR: Gender
RIDAGEYR: Age in years at screening
RIDRETH3: Race/Hispanic origin w/ NH Asian
RIDEXPRG: Pregnancy status at exam
BMXWT: Weight (kg)
BMXHT: Standing Height (cm)
BMXBMI: Body Mass Index (kg/m**2)
ALQ101: Had at least 12 alcohol drinks/1 yr.?
ALQ110: Had at least 12 alcohol drinks/lifetime?
ALQ120Q: How often drink alcohol over past 12 mos.
ALQ120U: # days drink alcohol per wk., mos., yr.
ALQ130: Avg # alcoholic drinks/day -past 12 mos.
ALQ141Q: # days have 4/5 drinks -past 12 mos.
ALQ141U: # days per week, month, year?
ALQ151: Ever have 4/5 or more drinks every day?
ALQ160: # days have 4/5 or more drinks in 2 hrs.
LBXGH: Glycohemoglobin (%)
URXP01: 1-Hydroxynaphthalene (ng/L)
URXP02: 2-Hydroxynaphthalene (ng/L)
URXP03: 3-Hydroxyfluorene (ng/L)
URXP04: 2-Hydroxyfluorene (ng/L)
URXP06: 1-Hydroxyphenanthrene (ng/L)
URXP10: 1-Hydroxypyrene (ng/L)
URXP25: 2 & 3-Hydroxyphenanthrene (ng/L)
LBXTC: Total Cholesterol (mg/dL)
LBDHDD: Direct HDL-Cholesterol (mg/dL)
LBXTR: Triglyceride (mg/dL)
LBDLDL: LDL-cholesterol (mg/dL)
LBXHSCRP: HS C-Reactive Protein (mg/L)
LBXSCR: Creatinine, refrigerated serum (mg/dL)
LBXMOPCT: Monocyte percent (%).

## Acknowledgements

The author thanks the NHANES for the dataset.

