## Supplemental tables for "Effects of environmental polycyclic aromatic hydrocarbon exposure and pro-Inflammatory activity on Type 2 Diabetes Mellitus"

**SUPPLEMENT SECTION**

Supplementary Table 1. Reference range table for urinary metabolites concentrations of polyaromatic hydrocarbons in males and females of US population normalized with creatinine value (Grainger, 2006).

| Analyte | Geometric mean (95% confidence level) 2009-10~ or 2013-14~ year creatinine corrected in ng/gm of creatinine |
| --- | --- |
|  | total |
| 1-hydroxynaphthalene | 2140 (1990-2320) |
| 2-hydroxynaphthalene | 3710 (3470-3970) |
| 2-hydroxyfluorene | 250 (237-264) |
| 3-hydroxyfluorene | 99.2 (93-106) |
| 1-hydroxyphenanthrene | 137 (132-143) |
| 2- & 3-hydroxyphenanthrene | 133 (127-140) |
| 1-hydroxypyrene | 125 (116-134) |

Footnote: Derived from NHANES data [Fourth National Report on Human Exposure to Environmental Chemicals Update (cdc.gov)](https://www.cdc.gov/exposurereport/pdf/FourthReport_UpdatedTables_Volume1_Jan2019-508.pdf) (Grainger et al., 2006).

Supplementary Table 2. Names and source location of the data used from the NHANES data set.

| Data type | Source | Variables |
| --- | --- | --- |
| Demographic data | [DEMO_I (cdc.gov)](https://wwwn.cdc.gov/Nchs/Nhanes/2015-2016/DEMO_I.htm) | Examination status, Age, Gender, Race, Weight and Pregnancy status |
| Examination data | [NHANES 2015-2016 Examination Data (cdc.gov)](https://wwwn.cdc.gov/nchs/nhanes/Search/DataPage.aspx?Component=Examination&CycleBeginYear=2015) | Blood pressure and Body measures |
| Questionnaire data | [NHANES 2015-2016 Questionnaire Data (cdc.gov)](https://wwwn.cdc.gov/nchs/nhanes/Search/DataPage.aspx?Component=Questionnaire&CycleBeginYear=2015) | Alcohol use, Diabetes status |
| Laboratory data | [NHANES 2015-2016 Laboratory Data (cdc.gov)](https://wwwn.cdc.gov/nchs/nhanes/Search/DataPage.aspx?Component=Laboratory&CycleBeginYear=2015) | Cholesterol, Triglycerides, Glycohoemoglobin, Hepatitis, Complete blood counts and PAH metabolites measurements |

Reference to the Supplement Section

Grainger, J., Huang, W., Patterson Jr, D. G., Turner, W. E., Pirkle, J., Caudill, S. P., . . . Sampson, E. J. (2006). Reference range levels of polycyclic aromatic hydrocarbons in the US population by measurement of urinary monohydroxy metabolites. *Environmental Research, 100*(3), 394-423.
